# Evaluation of the diagnostic value of YiDiXie^™^-SS, YiDiXie^™^-HS and YiDiXie^™^-D in breast cancer

**DOI:** 10.1101/2024.07.02.24309738

**Authors:** Yutong Wu, Huimei Zhou, Xutai Li, Chen Sun, Zhenjian Ge, Wenkang Chen, Yingqi Li, Shengjie Lin, Pengwu Zhang, Wuping Wang, Siwei Chen, Wei Li, Hui Hu, Xiaoling Liu, Yongqing Lai

**Affiliations:** Department of Urology, Peking University Shenzhen Hospital, Shenzhen, 518036; Institute of Urology, Shenzhen Peking University-The Hong Kong University of Science and Technology Medical Center, Shenzhen, 518036; Shantou University Medical College, Shantou, Guangdong 515041; The Fifth Clinical Medical College of Anhui Medical University, Hefei 230032; Shenzhen University Health Science Center, Shenzhen, China 518055; Shenzhen KeRuiDa Health Technology Co., Ltd., Shenzhen, 518071; Department of Specialized Medicine, Peking University Shenzhen Hospital, Shenzhen, 518036; Department of Breast and Thyroid Surgery, Peking University Shenzhen Hospital, Shenzhen, 518036

## Abstract

**Background:** Breast cancer is a serious threat to women’s health and breast cancer screening is of great importance. Breast ultrasound or mammography is widely used for screening or diagnosis of breast tumors, However, false-positive breast ultrasound or mammogram results can lead to misdiagnosis and wrong puncture biopsy, while false-negative breast ultrasound or mammogram results can lead to missed diagnosis and delayed treatment. There is an urgent need to find a convenient, cost-effective and noninvasive method to reduce the false-positive rate and the false-negative rate of breast ultrasound or mammography. The aim of this study was to evaluate the diagnostic value of YiDiXie™-SS, YiDiXie™-HS and YiDiXie™-D in breast cancer.

**Patients and methods:** 816 subjects (malignant group, n=778; benign tumor group, n=38) were finally included in this study. The remaining serum samples were collected and tested by YiDiXie™ all-cancer detection kit. The sensitivity and specificity of YiDiXie™ tests were evaluated respectively.

**Results:** The sensitivity of YiDiXie™-SS was 97.8% (95% CI: 96.5% - 98.6%) and its specificity was 63.2% (95% CI: 47.3% - 76.6%). This means that YiDiXie™-SS has an extremely high sensitivity and relatively high specificity in breast tumors.YiDiXie™-HS has a sensitivity of 84.4% (95% CI: 81.7% - 86.8%) and a specificity of 86.8% (95% CI: 72.7% - 94.2%). This means that YiDiXie™-HS has high sensitivity and specificity in breast tumors.YiDiXie™-HS has a sensitivity of 73.8% (95% CI: 70.6% - 76.7%) and a specificity of 94.7% (95% CI: 82.7% - 99.1%). This means that YiDiXie™-D has relatively high sensitivity and very high specificity in breast tumors. The sensitivity of YiDiXie™-SS in ultrasound and mammography-positive patients was 97.8% (95% CI: 96.5% - 98.7%), 97.6% (95% CI: 95.8% - 98.6%), and the specificity was 63.6% (95% CI. 46.6% - 77.8%), 58.3% (95% CI: 32.0% - 80.7%), respectively. This means that the application of YiDiXie™-SS reduced ultrasound and mammography false-positive rates by 63.6% (95% CI: 46.6% - 77.8%) and 58.3% (95% CI: 32.0% - 80.7%), respectively, with essentially no increase in the leakage of malignant tumors. The sensitivity of YiDiXie™-HS in ultrasound and mammography-negative patients was 84.8% (95% CI: 69.1% - 93.9%), 85.7% (95% CI: 76.7% - 91.6%), and the specificity was 60.0% (95% CI. 23.1% - 92.9%), 80.0% (95% CI: 37.6% - 99.0%), respectively. This means that the application of YiDiXie™-HS reduced the false-negative rates of ultrasound and mammography by 84.8% (95% CI: 69.1% - 93.9%), 85.7% (95% CI: 76.7% - 91.6%), respectively. The sensitivity of YiDiXie™-D in ultrasound and mammography-positive patients was 74.0% (95% CI: 70.8% - 77.0%), 76.9% (95% CI: 73.0% - 80.4%), and the specificity was 93.9% (95% CI. 80.4% - 98.9%), 91.7% (95% CI: 64.6% - 99.6%), respectively. This means that YiDiXie™-D reduced the false positive rate of ultrasound and mammography by 93.9% (95% CI: 80.4% - 98.9%), 91.7% (95% CI: 64.6% - 99.6%), respectively. The sensitivity of YiDiXie™ -D in ultrasound and mammography-negative patients was 72.7% (95% CI: 55.8% - 84.9%), 75.0% (95% CI: 64.8% - 83.0%), and its specificity was 100% (95% CI. 56.6% - 100%), 100% (95% CI: 56.6% - 100%), respectively. This means that YiDiXie™-D reduces the false negative rate of ultrasound and mammography by 72.7% (95% CI: 55.8% - 84.9%) and 75.0% (95% CI: 64.8% - 83.0%), respectively, while maintaining a high specificity.

**Conclusion:** YiDiXie™-SS has extremely high sensitivity and relatively high specificity in breast tumors.YiDiXie™-HS has high sensitivity and high specificity in breast tumors.YiDiXie™-D has relatively high sensitivity and extremely high specificity in breast tumors.YiDiXie™-SS significantly reduces the rate of false positives by breast ultrasound or mammography with essentially no increase in delayed treatment of breast cancer. YiDiXie™-HS significantly reduces the rate of false negatives on breast ultrasound or mammograms.YiDiXie™-D significantly reduces the rate of false positives on breast ultrasound or mammograms or significantly reduces the rate of false negatives while maintaining a high level of specificity. YiDiXie™ tests has significant diagnostic value in breast cancer and is expected to solve the problems of “high false positive rate” and “high false negative rate” of breast ultrasound or mammography.

## INTRODUCTION

Breast cancer is the most common malignant tumor in women. The latest data show that in 2022, there will be 2,308,897 new cases of breast cancer globally, ranking 2nd in the global rate of new malignant tumors; there will be 665,684 new deaths, ranking 4th in the global mortality rate of malignant tumors^1^; and the incidence rate of breast cancer in 2022 has increased by 2% compared with that in 2020^1,2^, and the prevalence continues to increase year by year. Currently, breast cancer is treated with a combination of surgical treatment, endocrine therapy, and targeted therapy depending on the tumor type^3^. In an epidemiological survey in the United States that included 11 common malignant tumors and the economic burden of disease, breast cancer topped the list with $39 billion in annual treatment expenditures^4^. The 5-year survival rate for stage I breast cancer patients in the United States from 2009-2015 was upwards of ninety percent, compared to only 27 percent for stage IV^5^. Breast cancer screening has been shown to improve cure rates^6-8^. Therefore, breast cancer screening can significantly improve the prognosis of breast cancer and reduce the financial burden of breast cancer patients. Therefore, breast cancer is a serious threat to women’s health and breast cancer screening is of great importance.

Breast ltrasound or mammography is widely used in the screening or diagnosis of breast tumors, but breast ultrasound or mammography can produce a large number of false-positive results. According to the ultrasound BI-RADS grading system developed by the American College of Radiology (ACR), a diagnostic result in BI-RADS categories 4-5 is positive and requires histopathologic examination for final diagnosis^9^. Category 4 lesions have a wide range of malignancy risk spanning (2%-95%) and immediate pathologic biopsy is recommended^9^. Whereas the majority of biopsies of these 4a lesions are negative, the false-positive rate can be as high as 90%-98%^9^. False-positive results of breast ultrasound or mammography imply misdiagnosis of benign disease as malignancy. As breast ultrasound or mammography is positive, puncture biopsy is usually taken to obtain tissue specimens for pathologic diagnosis, from which the next step in the treatment plan is developed^10^. As a result, patients with false-positive breast ultrasound or mammography will have to bear the negative consequences of unnecessary puncture biopsy in terms of mental anguish, expensive examination costs, and physical injury. Therefore, there is an urgent need to find a convenient, cost-effective and non-invasive diagnostic method to reduce the false positive rate of breast ultrasound or mammography.

On the other hand, breast ultrasound or mammography can produce a large number of false-positive results. According to the ultrasound BI-RADS grading system developed by the American College of Radiology (ACR), a diagnosis of a BI-RADS ≤ 3 category is considered negative^9^. When a breast ultrasound or mammogram is negative, patients are usually taken for observation and regular follow-up ^10^. A false negative breast ultrasound or molybdenum result implies a missed diagnosis of breast cancer, which will likely lead to a delay in treatment, progression of the malignancy, and possibly even development of advanced stages. Patients will thus have to bear the adverse consequences of poor prognosis, high treatment costs, poor quality of life, and short survival. Therefore, there is an urgent need to find a convenient, cost-effective and non-invasive diagnostic method to reduce the false-negative rate of breast ultrasound or molybdenum target.

Based on the detection of miRNAs in serum, Shenzhen KeRuiDa Health Technology Co., Ltd. has developed “YiDiXie ™ all-cancer test” (hereinafter referred to as YiDiXie™ tests)^11^. With only 200 milliliters of whole blood or 100 milliliters of serum, the test can detect multiple cancer types, enabling detection of cancer at home^11^. YiDiXie™ tests consists of three independent tests: YiDiXie™ -HS, YiDiXie™-SS and YiDiXie™-D^11^.

The purpose of this study is to evaluate the diagnostic value of YiDiXie™ tests in breast cancer.

## PATIENTS AND METHODS

### Study design

This work is part of the sub-study “Evaluating the diagnostic value of YiDiXie™ tests in multiple tumors” of the SZ-PILOT study (ChiCTR2200066840).

SZ-PILOT is a prospective, observational, single-center study (ChiCTR2200066840). At the time of admission or physical examination, subjects who signed a pan-informed permission form for the donation of their remaining samples were considered included, and 0.5 milliliter of their remaining serum samples was taken for this investigation.

The participants and investigators in this study were blinded. The laboratory professionals who administered YiDiXie ™ tests and the technicians of KeRuiDa Co. who determined the test findings were unaware of the participants’ clinical information. The clinical specialists evaluating the individuals’ clinical information were ignorant of the results of YiDiXie™ tests.

The Ethics Committee of Shenzhen Hospital of Peking University approved the study, which was carried out in accordance with the International Conference on Harmonization (ICH) Code of Practice for the Quality Management of Pharmaceutical Clinical Trials and the Declaration of Helsinki.

### Participants

According to the ultrasound BI-RADS grading system developed by the American College of Radiology (ACR), a diagnostic result in BI-RADS categories 4-5 is positive^9^.

Patients with breast ultrasound findings of BI-RADS category 4 or 5 were included in this study. The two groups were enrolled independently, and subjects who satisfied the inclusion criteria were added one after the other.

This study initially included hospitalized patients with “suspected (solid or hematological) malignancy” who signed a pan-informed agreement to donate the remaining samples. Subjects having a postoperative pathologic diagnostic of “breast cancer” were placed in the prostate cancer group, whereas those with a postoperative pathologic diagnosis of benign disease were put in the benign group. Participants who had unclear pathologic results were excluded from the research. Some of these malignant group were used in the previous study of our group^11^.

The study eliminated subjects who did not pass the serum sample quality test prior to YiDiXie ™ tests. For further information on enrollment and exclusion, please see the subject group’s earlier article^11^.

### Sample collection, processing

The serum samples utilized in this investigation were obtained from serum leftover from a routine medical consultation, eliminating the need for extra blood sampling. Approximately 0.5 ml of serum was collected from the remaining serum of subjects in the Medical Laboratory and stored at -80? for subsequent use in YiDiXie™ tests.

### YiDiXie™ tests

YiDiXie™ tests is done using the YiDiXie™ all-cancer detection kit. YiDiXie™ tests is an in vitro diagnostic kit developed and manufactured by Shenzhen KeRuiDa Health Technology Co. Ltd.. It determines whether a subject has cancer by looking for the expression levels of several dozen miRNA biomarkers in their serum. It maintains the specificity and increases the sensitivity of a wide range of malignancies by integrating these independent assays in a contemporaneous testing format and predefining suitable criteria for each miRNA biomarker to guarantee that each miRNA marker is highly specific.

YiDiXie™ tests consists of three tests with very different characteristics: YiDiXie ™ -Highly Sensitive (YiDiXie ™ -HS), YiDiXie ™ -Super Sensitive (YiDiXie ™ -SS) and YiDiXie ™ -Diagnosis(YiDiXie ™ -D). The development of YiDiXie™-HS took specificity and sensitivity into consideration. The number of miRNA testing was greatly enhanced with YiDiXie™-SS in order to obtain very high sensitivity for all clinical stages of all malignant tumor types. YiDiXie ™ -D dramatically increases the diagnostic threshold of a single miRNA test to achieve very high specificity (very low misdiagnosis rate) for all malignancy types.

Perform YiDiXie ™ tests according to the instructions for the YiDiXie™ all-cancer detection kit. Refer to our prior article for detailed procedures^11^.

The raw results were analyzed by the laboratory technicians of KeRuiDa Co., and the results of YiDiXie™ tests were determined to be “positive” or “negative”.

### Diagnosis of breast ultrasound and mammography

The diagnosis of breast ultrasound and mammography was determined based on the BI-RADS grade of the included cases that had a BI-RADS grade. A BI-RADS grade of 4 or 5 is considered “positive” and a BI-RADS grade of ≤3 is considered “negative”^9^.If the included cases did not have a BI-RADS grade, the results were judged as “positive” or “negative” according to their diagnostic conclusion.

If the diagnostic conclusion was positive, more positive, or tended to be malignant, the result was judged as “positive”. If the diagnosis is positive, more positive or inclined to benign disease, or if the diagnosis is ambiguous, the test result is judged as “negative”.

### Clinical data collection

For this study, clinical, pathological, laboratory, and imaging data were retrieved from the subjects’ hospitalized medical records or physical examination reports. The clinical staging was evaluated by trained clinicians according to the AJCC staging manual (7th or 8th edition)^12,13^.

### Statistical analyses

Descriptive statistics were reported for demographic and baseline characteristics. Number and percentage of subjects in each category were calculated for categorical variables, and minimum and maximum values were calculated for total number of subjects (n), mean, standard deviation (SD) or standard error (SE), median, first quartile (Q1), third quartile (Q3), and minimum and maximum values for continuous variables. Wilson (score) method was used to calculate 95% confidence intervals (CI) for multiple indicators.

## RESULTS

### Participant disposition

816 study participants were involved in this research (n = 778 cases for the malignant group and 38 cases for the benign group). The 816 participants’ clinical and demographic details are listed in Table 1.

**Table 1.** Participants’ demographic and clinical manifestation.

|  | Malignant (n = 778) | Benign (n = 38) | Total (N = 816) |
| --- | --- | --- | --- |
| Age, years |  |  |  |
| Mean (SD) | 49.5 (12.00) | 43.3 (11.78) | 49.2 (12.06) |
| Median (Q1,Q3) | 48 (41, 57) | 42 (35, 51) | 48 (41, 57) |
| Min, max | 22, 88 | 20, 71 | 20, 88 |
| Age, group, n (%) |  |  |  |
| <50 | 426 (54.8) | 28 (73.7) | 454 (55.6) |
| ≥ 50 | 352 (45.2) | 10 (26.3) | 362 (44.4) |
| <65 | 684 (87.9) | 36 (94.7) | 720 (88.2) |
| ≥ 65 | 94 (12.1) | 2 (5.3) | 96 (11.8) |
| Body mass index(kg/m2) |  |  |  |
| n | 763 | 37 | 800 |
| Mean (SD) | 23.1 (3.02) | 22.9 (3.87) | 23.1 (3.06) |
| Median (Q1,Q3) | 22.8 (21.1, 24.8) | 22.3 (20.7, 26.0) | 22.8 (21.0, 24.8) |
| Min, max | 14.6 35.9 | 16.3 32.0 | 14.6 35.9 |
| Body mass index category, n (%) |  |  |  |
| Underweight | 31 (4.0) | 5 (13.2) | 36 (4.4) |
| Benign | 475 (61.1) | 21 (55.3) | 496 (60.8) |
| Overweight | 206 (26.5) | 7 (18.4) | 213 (26.1) |
| Obese | 51 (6.6) | 4 (10.5) | 55 (6.7) |
| Missing | 15 (1.9) | 1 (2.6) | 16 (2.0) |
| AJCC clinical stage |  |  |  |
| Stage 0 | 74 (9.5) |  | 74 (9.5) |
| Stage I | 288 (37.0) |  | 288 (37.0) |
| Stage II | 329 (42.3) |  | 329 (42.3) |
| Stage III | 87 (11.2) |  | 87 (11.2) |

Q1,Q3, first quartile, third quartile; SD, standard deviation.

In terms of clinical and demographic traits, the two study subject groups were similar (Table 1). The mean (standard deviation) age was 49.2 (12.06) years.

### Diagnostic Performance of YiDiXie™-SS

As shown in Table 2, the sensitivity of YiDiXie™-SS was 97.8% (95% CI: 96.5% - 98.6%) and its specificity was 63.2% (95% CI: 47.3% - 76.6%). This means that YiDiXie™-SS has very high sensitivity and relatively high specificity in breast tumors.

**Table 2.** The performance of YiDiXie™-SS.

| Table 2. The performance of YiDiXie™-SS |  |  |  |
| --- | --- | --- | --- |
|  | Malignant | Benign | Total |
|  | 778 | 38 | 816 |
| Positive | 761 | 14 | 775 |
| Negative | 17 | 24 | 41 |
| $SEN = 761/778 = 97.8\%(96.5\% - 98.6\%)$ $SPE = 24/38 = 63.2\%(47.3\% - 76.6\%)$ | | | |
| SEN, Sensitivity. SPE, Specificity. Two-sided 95%Wilson confidence intervals were calculated. |  |  |  |

### Diagnostic performance of YiDiXie™-HS

As shown in Table 3, the sensitivity of YiDiXie™-HS was 84.4% (95% CI: 81.7% - 86.8%) and its specificity was 86.8% (95% CI: 72.7% - 94.2%). This means that YiDiXie™-HS has high sensitivity and high specificity in breast tumors.

**Table 3.** The performance of YiDiXie™-HS.

| Table 3. The performance of YiDiXie™-HS |  |  |  |
| --- | --- | --- | --- |
|  | Malignant | Benign | Total |
|  | 778 | 38 | 816 |
| Positive | 657 | 5 | 662 |
| Negative | 121 | 33 | 154 |
| $SEN = 657/778 = 84.4\%(81.7\% - 86.8\%)$ $SPE = 33/38 = 86.8\%(72.7\% - 94.2\%)$ | | | |
| SEN, Sensitivity. SPE, Specificity. Two-sided 95%Wilson confidence intervals were calculated. |  |  |  |

### Diagnostic performance of YiDiXie™-D

As shown in Table 4, the sensitivity of YiDiXie™-D was 73.8% (95% CI: 70.6% - 76.7%) and its specificity was 94.7% (95% CI: 82.7% - 99.1%). This means that YiDiXie ™ -D has relatively high sensitivity and very high specificity in breast tumors.

**Table 4.**
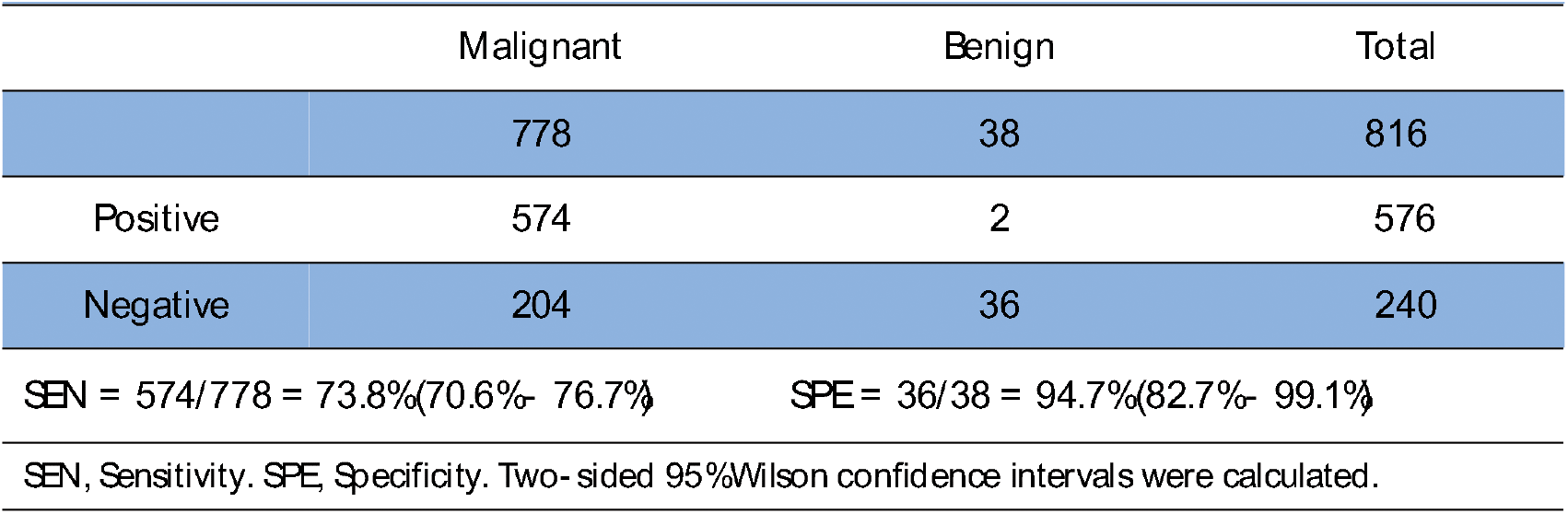
Performance of YiDiXie™-D.

| Table 4. Performance of YiDiXie™ D |  |  |  |
| --- | --- | --- | --- |
|  | Malignant | Benign | Total |
|  | 778 | 38 | 816 |
| Positive | 574 | 2 | 576 |
| Negative | 204 | 36 | 240 |
| SEN = 574/778 = 73.8%(70.6%- 76.7%) SPE = 36/38 = 94.7%(82.7%- 99.1%) |  |  |  |
| SEN, Sensitivity. SPE, Specificity. Two-sided 95%Wilson confidence intervals were calculated. |  |  |  |

### Diagnostic performance of YiDiXie™-SS in breast ultrasound-positive patients

In order to solve the problem of high false-positive rate of breast ultrasound, YiDiXie™ -SS was applied to breast ultrasound-positive patients.

As shown in Table 5, the sensitivity of YiDiXie ™ -SS in breast ultrasound-positive patients was 97.8% (95% CI: 96.5% - 98.7%), while the specificity was 63.6% (95% CI: 46.6% - 77.8%). This means that the application of YiDiXie ™ -SS reduces the rate of breast ultrasound false positives by 63.6% (95% CI: 46.6% - 77.8%) with essentially no increase in malignant tumor underdiagnosis.

**Table 5.** Performance of different Items.

| Items | Total N | Positive | Sensitivity %(95%CI)<sup>a</sup> | Total N | Negative | Specificity %(95%CI)<sup>b</sup> |
| --- | --- | --- | --- | --- | --- | --- |
| YiDiXie™ SSin US- positive | 743 | 727 | 97.8%(96.5%- 98.7%) | 33 | 21 | 63.6%(46.6%- 77.8%) |
| YiDiXie™ HSin US- negative | 33 | 28 | 84.8%(69.1%- 93.9%) | 5 | 3 | 60.0%(23.1%- 92.9%) |
| YiDiXie™ D in US- positive | 743 | 550 | 74.0%(70.8%- 77.0%) | 33 | 31 | 93.9%(80.4%- 98.9%) |
| YiDiXie™ D in US- negative | 33 | 24 | 72.7%(55.8%- 84.9%) | 5 | 5 | 100%(56.6%- 100%) |
| YiDiXie™ SSin Xray- positive | 497 | 485 | 97.6%(95.8%- 98.6%) | 12 | 7 | 58.3%(32.0%- 80.7%) |
| YiDiXie™ HSin X ray- negative | 84 | 72 | 85.7%(76.7%- 91.6%) | 5 | 4 | 80.0%(37.6%- 99.0%) |
| YiDiXie™ D in Xray- positive | 497 | 382 | 76.9%(73.0%- 80.4%) | 12 | 11 | 91.7%(64.6%- 99.6%) |
| YiDiXie™ D in Xray- negative | 84 | 63 | 75.0%(64.8%- 83.0%) | 5 | 5 | 100%(56.6%- 100%) |

CI, confidence interval. <sup>a,b</sup> Two- sided 95%Wilson confidence intervals were calculated.

### Diagnostic performance of YiDiXie™-HS in breast ultrasound-negative patients

In order to solve the problem of high false-negative rate of breast ultrasound, YiDiXie ™-HS was applied to breast ultrasound-negative patients.

As shown in Table 5, the sensitivity of YiDiXie ™ -HS in breast ultrasound-negative patients was 84.8% (95% CI: 60.1% - 93.3%), and the specificity was 60.0% (95% CI: 23.1% - 92.9%). This means that the application of YiDiXie ™ -HS reduces the rate of breast ultrasound false negatives by 84.8% (95% CI: 60.1% - 93.3%).

### Diagnostic performance of YiDiXie™-D in breast ultrasound-positive patients

To further reduce the rate of breast ultrasound false positives, YiDiXie™-D, which has a relatively high sensitivity and very high specificity, was therefore applied.

As shown in Table 5, the sensitivity of YiDiXie™-D in breast ultrasound-positive patients was 74.0% (95% CI: 70.8% - 77.0%) and its specificity was 93.9% (95% CI: 80.4% - 98.9%). This means that YiDiXie™-D reduces the false positive rate of breast ultrasound by 93.9% (95% CI: 80.4% - 98.9%).

### Diagnostic performance of YiDiXie™-D in breast ultrasound-negative patients

In order to reduce the breast ultrasound false-negative rate while maintaining high specificity, YiDiXie™-D, which has relatively high sensitivity and very high specificity, was applied.

As shown in Table 5, the sensitivity of YiDiXie ™ -D in breast ultrasound-negative patients was 72.7% (95% CI: 55.8% - 84.9%) and its specificity was 100% (95% CI: 56.6% - 100%). This means that YiDiXie ™ -D reduces the false-negative rate of breast ultrasound by 72.7% (95% CI: 55.8% - 84.9%) while maintaining high specificity.

### Diagnostic performance of YiDiXie™-SS in mammography-positive patients

In order to solve the problem of high false-positive rate of mammography, YiDiXie™ -SS was applied to mammography-positive patients.

As shown in Table 5, the sensitivity of YiDiXie™-SS was 97.6% (95% CI: 95.8% - 98.6%) and the specificity was 58.3% (95% CI: 32.0% - 80.7%). This means that the application of YiDiXie™-SS reduces the false-positive rate of mammograms by 58.3% (95% CI: 32.0% - 80.7%) with essentially no increase in malignant tumor underdiagnosis.

### Diagnostic performance of YiDiXie™-HS in mammography-negative patients

In order to solve the challenge of high false-negative rate of mammography, YiDiXie™ -HS was applied to mammography-negative patients.

As shown in Table 5, the sensitivity of YiDiXie™-HS in mammography-negative patients was 85.7% (95% CI: 76.7% - 91.6%), specificity was 80.0% (95% CI: 37.6% - 99.0%). This means that the application of YiDiXie™-HS reduces the false-negative mammogram rate by 85.7% (95% CI: 76.7% - 91.6%).

### Diagnostic performance of YiDiXie™-D in mammography-positive patients

In order to further reduce the rate of false-positive mammograms, YiDiXie™-D, which has a relatively high sensitivity and very high specificity, was therefore applied.

As shown in Table 5, the sensitivity of YiDiXie ™-D in mammography-positive patients was 76.9% (95% CI: 73.0% - 80.4%) and its specificity was 91.7% (95% CI: 64.6% - 99.6%). This means that YiDiXie™-D reduces the false positive rate of mammograms by 91.7% (95% CI: 64.6% - 99.6%).

### Diagnostic performance of YiDiXie™-D in mammography-negative patients

In order to reduce the false-negative rate of mammograms while maintaining a high degree of specificity, YiDiXie™-D, which has a relatively high sensitivity and very high degree of specificity, was applied.

As shown in Table 5, the sensitivity of YiDiXie™-D in mammography-negative patients was 75.0% (95% CI: 64.8% - 83.0%) and its specificity was 100% (95% CI: 56.6% - 100%). This means that YiDiXie ™-D reduces the false-negative rate of mammograms by 75.0% (95% CI: 64.8% - 83.0%) while maintaining high specificity.

## DISCUSSION

### Clinical significance of YiDiXie™-SS in breast ultrasound or mammography-positive patients

There are 3 very different tests in the 1-Drop ™ Test: YiDiXie ™ -HS, YiDiXie ™ -SS, and YiDiXie ™ -D. YiDiXie ™ -HS has both high sensitivity and high specificity. YiDiXie™-SS has very high sensitivity for all types of malignant tumors, but slightly lower specificity. YiDiXie™-D has very high specificity for all types of malignant tumors, but relatively low sensitivity.

For breast ultrasound or mammography-positive patients, both sensitivity and specificity of further diagnostic methods are important. On one hand, the sensitivity is critical. A lower sensitivity means a higher rate of false negatives. When the results of this diagnostic method are negative, the diagnosis usually ends for that patient. A higher false-negative rate means that more malignant tumors are missed, which will lead to a delay in their treatment, progression of the malignant tumor, and possibly even development of advanced stages. The patients will thus be obliged to bear the adverse consequences of poor prognosis, poor quality of life, and high cost of treatment.

On the other hand, the specificity is very important. Lower specificity means a higher rate of false positives. When the diagnosis is positive, breast tumors are usually biopsied by puncture biopsy. A higher false positive rate means that more cases of benign tumors undergo puncture biopsy. It definitely leads to a significant increase in emotional distress, costly procedures or tests, physical injuries, and other negative consequences for the patient.

Consequently, there is a trade-off between “fewer malignant tumors missed” and “fewer benign tumors misdiagnosed” when it comes to sensitivity and specificity. When non-breast cancer cases is mistakenly identified as malignant tumor, aspiration biopsy is typically performed instead of surgical resection. Therefore, false-positive breast ultrasound does not lead to serious consequences of organ loss. Therefore, for breast ultrasound-positive patients, “fewer missed diagnoses of malignant tumors” is considerably more essential than “fewer misdiagnoses of benign tumors.” Therefore, YiDiXie ™ -SS was chosen for reducing the false-positive rates of breast ultrasound rather than YiDiXie™-HS or YiDiXie™-D.

As shown in Tables 5, YiDiXie™-SS had a sensitivity of 97.8% (95% CI: 96.5% - 98.7%), 97.6% (95% CI: 95.8% - 98.6%), and specificity of 63.6% (95% CI: 46.6% - 77.8%), 58.3% (95% CI: 32.6% - 77.8%) in breast ultrasound or mammography-positive patients. These results indicate that YiDiXie™-SS reduces false positives on breast ultrasound or mammography by 63.6% (95% CI: 46.6% - 77.8%), 58.3% (95% CI: 32.0% - 80.7%), respectively, while maintaining sensitivity close to 100%.

As mentioned above, missed diagnosis of breast cancer means delayed treatment, while false positive results of breast ultrasound or mammography mean puncture biopsy. The above results imply that YiDiXie ™ -SS dramatically reduces the probability of false puncture biopsies of benign breast tumors with essentially no increase in malignant tumor underdiagnosis.

In other words, YiDiXie ™ -SS drastically decreases the mental suffering, expensive examination or surgery costs, physical injuries, and other adverse consequences for breast ultrasound or mammography-positive patients without basically increasing the delayed treatment of malignant tumors. Therefore, YiDiXie ™ -SS well fulfills the clinical demand and has important clinical significance and broad prospective application.

### Clinical significance of YiDiXie™-HS in breast ultrasound or mammography-negative patients

For breast ultrasound or mammography-negative patients, both the sensitivity and specificity of further diagnostic methods are of great importance. On the one hand, sensitivity is crucial. Higher false-negative rates mean that more malignancies are missed. On the other hand, specificity is crucial. Higher false-positive rates mean more benign tumors are misdiagnosed. In general, benign breast tumors misdiagnosed as malignant tumors usually undergo puncture biopsy rather than radical resection. Therefore, false-positive breast ultrasound does not lead to serious consequences of organ removal. Thus, for breast ultrasound or mammography-negative patients, “fewer malignant tumors missed” is more important than “fewer benign tumors misdiagnosed”.

In addition, the negative predictive value is higher in breast ultrasound or mammography-negative patients. In this way, both higher false-positive and false-negative rates can lead to significant harm. Therefore, YiDiXie™ -HS, with its high sensitivity and specificity, was chosen to reduce the breast ultrasound or mammography false-negative rate.

As shown in Tables 5, the sensitivity of Y iDiXie™-HS in breast ultrasound or mammography-negative patients was 84.8% (95% CI: 6 0.1% - 93.3%), 85.7% (95% CI: 76.7% - 91. 6%), and specificity was 60.0% (95% CI: 23.1% - 92.9%), 80.0% (95% CI: 37.6% - 99.0%). T his means that YiDiXie™-HS reduced false negatives on breast ultrasound or mammogram by 84.8% (95% CI: 60.1% - 93.3%), 85.7% (9 5% CI: 76.7% - 91.6%), respectively.

The above results imply that YiDiXie™-HS significantly reduces the probability of missing malignant tumors with false-negative results on breast ultrasound or mammography. Therefore, YiDiXie™-HS meets the clinical needs well and has important clinical significance and wide application prospects.

### Clinical significance of YiDiXie™-D

For patients with breast tumors, YiDiXie™-D, with its relatively high sensitivity and very high specificity, can be used to further reduce the rate of false positives or significantly reduce the rate of false negatives on breast ultrasound or mammograms while maintaining a high level of specificity.

As shown in Table 5, the sensitivity of Yi DiXie™-D in breast ultrasound or mammography-positive patients was 74.0% (95% CI: 70.8% - 77.0%), 76.9% (95% CI: 73.0% - 80.4%), a nd its specificity was 93.9% (95% CI: 80.4% - 98.9%), 91.7% (95% CI: 64.6% - 99.6%) res pectively; YiDiXie™-D had a sensitivity of 72.7% (95% CI: 55.8% - 84.9%) in breast ultrasound or mammography-negative patients. 55.8% - 84.9%), 75.0% (95% CI: 64.8% - 83.0%) in breast ultrasound or mammography-negative patients, respectively; and the specificity of YiDi Xie™-D was 100% (95% CI: 56.6% - 100%), 100% (95% CI: 56.6% - 100%). These results indicate that YiDiXie™-D reduced false positi ves by 93.9% (95% CI: 80.4% - 98.9%)?91.7% (95% CI: 64.6% - 99.6%), or 72.7% (95% CI: 55.8% - 84.9%)?75.0% (95% CI: 64.8% - 83. 0%) for breast ultrasound or mammography w hile maintaining high specificity.

The above results imply that YiDiXie™-D substantially reduces the probability of wrong puncture biopsy in breast tumors. Therefore, YiDiXie™-D well meets the clinical needs and has important clinical significance and wide application prospects.

### YiDiXie™ tests is expected to solve two problems of breast tumor

First, YiDiXie™-SS significantly reduces the risk of misdiagnosis of benign breast tumors as breast cancer. On the one hand, YiDiXie™-SS substantially reduces the probability of incorrect puncture biopsy of benign breast tumors with essentially no increase in breast cancer missed diagnosis. As shown in Tables 5, YiDiXie™-SS reduced the magnitude of false-positive breast ultrasound or molybdenum target by 63.6% (95% CI: 46.6% - 77.8%), and 58.3% (95% CI: 32.0% - 80.7%), respectively, with essentially no increase in breast cancer missed diagnosis. Thus, YiDiXie ™-SS significantly reduces the range of adverse outcomes associated with unnecessary mammocentesis biopsies with essentially no increase in delayed treatment of breast cancer.

On the other hand, YiDiXie ™-SS greatly eliminates nonessential workload for clinicians and contributes to the timely treatment of malignant tumor cases that would otherwise be delayed. When breast ultrasound or mammography is positive, patients usually undergo puncture biopsies. The availability of these biopsies is directly dependent on the number of clinicians. In many parts of the world, puncture biopsies are not performed until appointments have been made for months or even more than a year. It inevitably slows down the treatment of malignancy cases, and therefore it is not uncommon for breast tumor patients awaiting treatment to develop malignancy progression or even distant metastases.

As shown in Tables 5, YiDiXie™-SS reduced the magnitude of false-positive breast ultrasound or molybdenum target by 63.6% (95% CI: 46.6% - 77.8%), and 58.3% (95% CI: 32.0% - 80.7%), respectively, with essentially no increase in breast cancer missed diagnosis. As a result, YiDiXie™ -SS significantly relieves physician of unnecessary workload and facilitates the timely treatment of malignant tumors that would otherwise be delayed.

Second, YiDiXie™-HS significantly reduces the risk of breast cancer being missed. When a breast ultrasound or mammogram is negative, the possibility of breast cancer is usually ruled out for the time being. The high false-negative rate of breast ultrasound or mammography results in a large number of breast cancer patients delaying treatment. As shown in Tables 5, YiDiXie™-HS reduced false-negative breast ultrasound or mammography by 84.8% (95% CI: 60.1% - 93.3%), 85.7% (95% CI: 76.7% - 91.6%), respectively. As a result, YiDiXie ™ -HS significantly reduces the likelihood of missed diagnosis of malignant tumors due to false-negative results on breast ultrasound or mammography, and facilitates timely diagnosis and treatment of breast cancer patients who would otherwise be delayed in treatment.

Again, YiDiXie™-D is expected to further address the challenges of “ high false positive rate “ and “ high false negative rate “. Breast tumors are usually biopsied when malignancy is considered. However, in the case of smaller tumors, low BI-RADS scores, etc., false positives will lead to unnecessary puncture biopsies. Therefore these patients require extra caution before puncture biopsy. As shown in Table 5, YiDiXie ™ -D reduced breast ultrasound or mammogram false positives by 93.9% (95% CI: 80.4% - 98.9%)?91.7% (95% CI: 64.6% - 99.6%), respectively, or breast ultrasound or mammogram false negatives by 72.7% (95% CI: 55.8% - 84.9%), 75.0% (95% CI: 64.8% - 83.0%), respectively. Thus, YiDiXie ™ -D substantially reduces the probability of wrong puncture biopsy in breast tumors.

Final, YiDiXie ™ tests enable “just-in-time diagnosis” for breast tumor patients. On one hand, YiDiXie ™ tests only requires microscopic amounts of blood, allowing patients to complete the diagnostic process non-invasively without leaving their homes. A single YiDiXie ™ test needs only 20 microliters of serum, which is approximately the same amount as one drop of whole blood (one drop of whole blood is about 50 microliters, which yields 20-25 microliters of serum). Given the pre-test sample quality assessment and 2-3 repetitions, 0.2 mL of whole blood is enough to complete YiDiXie™ tests. The 0.2 mL of finger blood can be collected at home by the average patient using a finger blood collection needle, instead of requiring venous blood collection by medical personnel, allowing the patient to complete the diagnostic process non-invasively without having to leave their home. On the other hand, the diagnostic capacity of YiDiXie ™ tests is almost limitless. Figure 1 shows the basic flow chart of YiDiXie™ tests, which shows that YiDiXie ™ tests does not require both a doctor and medical equipment, and does not require medical personnel to collect blood.

Thus, YiDiXie ™ tests are absolutely independent of the number of clinicians and healthcare organizations, and has a nearly unlimited testing capacity. Therefore, YiDiXie™ tests enables “just-in-time” diagnosis of breast ultrasound-positive patients without the need for patients to wait anxiously for an appointment.

In short, YiDiXie ™ tests have important diagnostic value in breast cancer, and are expected to solve the two difficult problems of “too high false-positive rate of breast ultrasound or mammography” and “too high false-negative rate of breast ultrasound or mammography” in breast tumors.

### Limitations of the study

Firstly, this study had a small number of cases and a clinical study with a larger sample size is needed for further evaluation in the future.

Secondly, this was an inpatient malignant tumor case-benign tumor control study, and future cohort studies of breast tumor patients are needed for further evaluation.

Finally, the current study was a single-center study, which may have led to some degree of bias in the results of this study. A multicenter study is needed for further evaluation in the future.

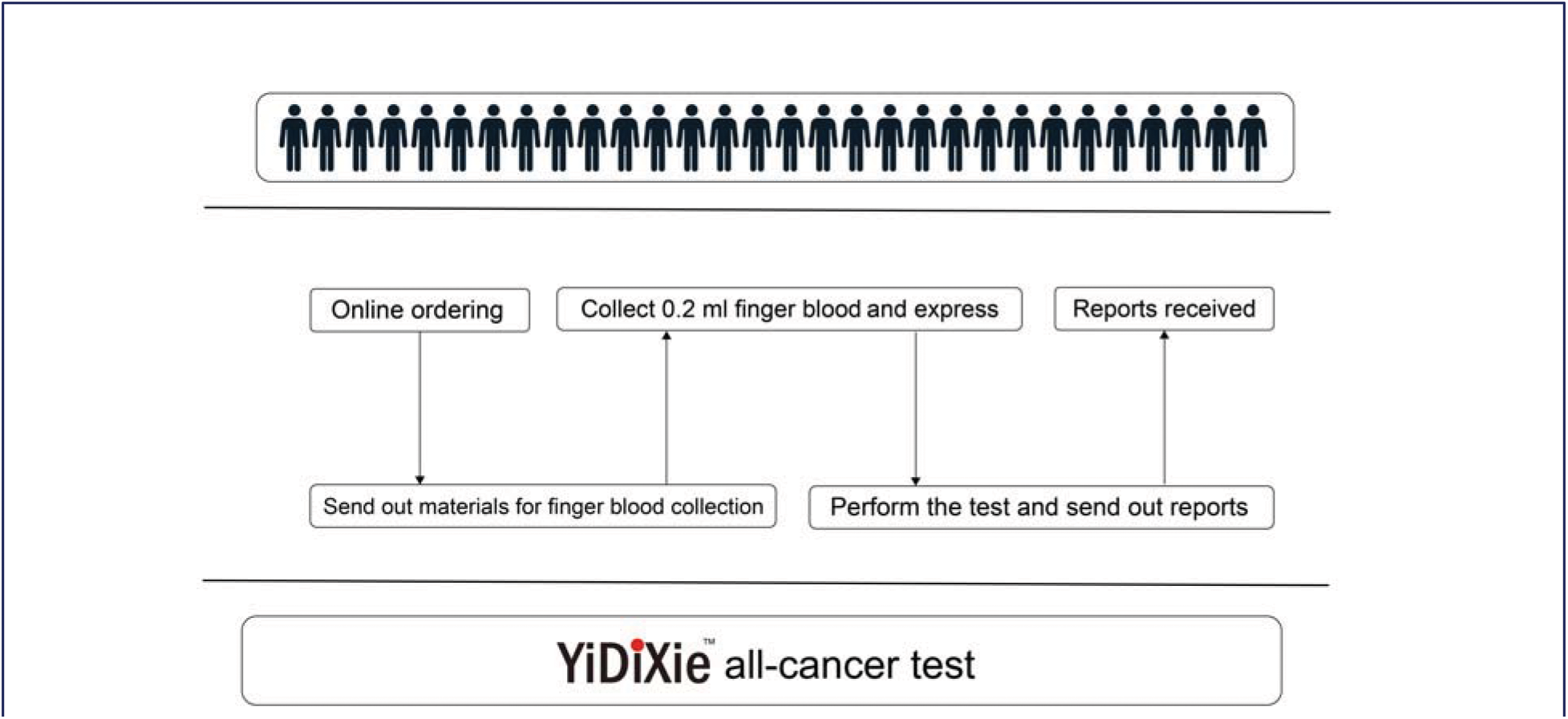

## CONCLUSION

YiDiXie™-SS has extremely high sensitivity and relatively high specificity in breast tumors.YiDiXie™-HS has high sensitivity and high specificity in breast tumors.YiDiXie ™ -D has relatively high sensitivity and extremely high specificity in breast tumors.YiDiXie ™ -SS significantly reduces the rate of false positives by breast ultrasound or mammography with essentially no increase in delayed treatment of breast cancer. YiDiXie™-HS significantly reduces the rate of false negatives on breast ultrasound or mammograms.YiDiXie™-D significantly reduces the rate of false positives on breast ultrasound or mammograms or significantly reduces the rate of false negatives while maintaining a high level of specificity. YiDiXie ™ tests has significant diagnostic value in breast cancer and is expected to solve the problems of “high false positive rate” and “high false negative rate” of breast ultrasound or mammography.

## Data Availability

All data produced in the present study are contained in the manuscript.

## FUNDING

This study was supported by Shenzhen High-level Hospital Construction Fund, Clinical Research Project of Peking University Shenzhen Hospital (LCYJ2020002, LCYJ2020015, LCYJ2020020, LCYJ2017001).

## Notes

### Competing Interest Statement

The authors have declared no competing interest.

### Clinical Trial

ChiCTR2200066840

### Funding Statement

This work was supported by Shenzhen High-level Hospital Construction Fund, Clinical Research Project of Peking University Shenzhen Hospital (LCYJ2020002, LCYJ2020015, LCYJ2020020, LCYJ2017001).

### Author Declarations

Ethics committee of Peking University Shenzhen Hospital gave ethical approval for this work.

### Summary of Updates

The results were updated. Most tabels and results were reviesed.

